# Population-based estimates of post-acute sequelae of SARS-CoV-2 infection (PASC) prevalence and characteristics: A cross-sectional study

**DOI:** 10.1101/2021.03.08.21252905

**Authors:** Jana L. Hirschtick, Andrea R. Titus, Elizabeth Slocum, Laura E. Power, Robert E. Hirschtick, Michael R. Elliott, Patricia McKane, Nancy L. Fleischer

## Abstract

**Importance:** Emerging evidence suggests many people have persistent symptoms after acute COVID-19 illness.

**Objective:** To estimate the prevalence and correlates of persistent COVID-19 symptoms 30 and 60 days post onset using a population-based sample.

**Design & Setting:** The Michigan COVID-19 Recovery Surveillance Study is a population-based cross-sectional survey of a probability sample of adults with confirmed COVID-19 in the Michigan Disease Surveillance System (MDSS). Respondents completed a survey online or via telephone in English, Spanish, or Arabic between June - December 2020.

**Participants:** Living non-institutionalized adults (aged 18+) in MDSS with COVID-19 onset through mid-April 2020 were eligible for selection (n=28,000). Among 2,000 adults selected, 629 completed the survey. We excluded 79 cases during data collection due to ineligibility, 6 asymptomatic cases, 7 proxy reports, and 24 cases missing outcome data, resulting in a sample size of 593. The sample was predominantly female (56.1%), aged 45 and older (68.2%), and Non-Hispanic White (46.3%) or Black (34.8%).

**Exposures:** Demographic (age, sex, race/ethnicity, and annual household income) and clinical factors (smoking status, body mass index, diagnosed comorbidities, and illness severity).

**Main outcomes and Measures:** We defined post-acute sequelae of SARS-CoV-2 infection (PASC) as persistent symptoms 30+ days (30-day COVID-19) or 60+ days (60-day COVID-19) post COVID-19 onset.

**Results:** 30- and 60-day COVID-19 were highly prevalent (52.5% and 35.0%), even among respondents reporting mild symptoms (29.2% and 24.5%) and non-hospitalized respondents (43.7% and 26.9%, respectively). Low income was statistically significantly associated with 30-day COVID-19 in adjusted models. Respondents reporting very severe (vs. mild) symptoms had 2.25 times higher prevalence of 30-day COVID-19 (Adjusted Prevalence Ratio [aPR] 2.25, 95% CI 1.46-3.46) and 1.71 times higher prevalence of 60-day COVID-19 (aPR 1.71, 95% 1.02-2.88). Hospitalized (vs. non-hospitalized) respondents had about 40% higher prevalence of both 30-day (aPR 1.37, 95% CI 1.12-1.69) and 60-day COVID-19 (aPR 1.40, 95% CI 1.02-1.93).

**Conclusions and Relevance:** PASC is highly prevalent among cases with severe initial symptoms, and, to a lesser extent, cases with mild and moderate symptoms.

## Introduction

The United States continues to be heavily impacted by the COVID-19 pandemic, with over 28 million confirmed cases and 517,000 deaths as of March 8, 2021.^1^ Emerging evidence indicates that a subset of people with COVID-19, commonly referred to as “long-haulers,” experience persistent symptoms for weeks or months after their COVID-19 diagnosis.^2-13^ Given the disproportionate number of cases in the US, and a global case tally over 117 million,^14^ long-term COVID-19 health effects for even a fraction of cases will have significant public health and economic implications.

We currently know very little about this prolonged course of COVID-19, also known as post-acute sequelae of SARS-CoV-2 infection (PASC). It appears to commonly manifest with symptoms of extreme fatigue,^3,6-11,13^ shortness of breath,^3,5,7,8,10^ persistent loss of taste or smell,^5,10,13^ and cognitive dysfunction.^5-7^ Due to the lack of a clear definition^15^ and very few population-based studies, prevalence estimates vary greatly, ranging from 13% of cases with persistent symptoms at least 4 weeks post diagnosis^11^ to 87% of hospitalized patients with symptoms an average of 60 days post symptom onset.^3^ Additionally, while some studies have shown an association between increasing age^11-13^ or female sex^11^ and PASC, others have not.^2,5^ Similarly, several studies report no association between comorbidities and risk of PASC,^2,5,13^ while others found increased risk among people with asthma,^11^ obesity,^12^ or a pre-existing psychiatric condition.^12^ Despite this lack of clarity, there is emerging evidence that severity of acute illness, measured by number of symptoms,^5,11,13^ severity score,^9^ or hospitalization,^11^ may increase the risk of PASC.

The existing literature base is narrow and includes very few population-based studies needed to provide accurate prevalence estimates. Furthermore, we do not have a clear understanding of who is at increased risk of experiencing PASC. Our objectives for this study are to 1) provide PASC prevalence estimates using a population-based sample of diagnosed COVID-19 cases in Michigan, and 2) assess demographic and clinical correlates of PASC.

## Methods

The University of Michigan institutional review board reviewed the study and determined it was exempt from ongoing IRB review due to the use of secondary de-identified data. This study followed the Strengthening the Reporting of Observational Studies in Epidemiology guidelines.

### MI CReSS Sample

The Michigan COVID-19 Recovery Surveillance Study (MI CReSS) is a population-based study of adults 18 and older with a PCR-confirmed SARS-CoV-2 test in the Michigan Disease Surveillance System (MDSS). All non-institutionalized adults with a valid phone number and zip code or county in the MDSS who were alive at the time the survey sample was drawn were eligible for selection (n=28,000). We drew a stratified, random sample of 2,000 adults with COVID-19 onset on or before April 15, 2020. COVID-19 onset was determined by self-reported symptom onset date when available (80% of sampled cases), followed by positive SARS-CoV-2 test date (19%), or date of referral to the Michigan Department of Health and Human Services (MDHHS; 1%). Sampling strata included 13 geographic areas in Michigan: six emergency preparedness regions^16^ each consisting of multiple counties, and six counties and one city (Detroit) in southeast Michigan, the epicenter of the initial outbreak in Michigan. Sampling weights were constructed using generalized regression estimators^17^ so that the weighted distribution of the sample matched the age and sex distribution by geographic region of the sampling frame.

We sent selected cases a recruitment letter in mid-June 2020, inviting them to complete a survey online or via telephone in English, Spanish, or Arabic. Of the 2,000 adults selected, 79 were subsequently excluded due to death (n=12); cognitive impairment (n=41); inability to complete the survey in the languages offered (n=12); or some other reason rendering them ineligible (n=14). Because our focus was self-reported length of recovery and symptoms, we also excluded asymptomatic cases (n=6), consistent with prior studies,^12^ and proxy reports due to cognitive ability or incapacitation (n=6). Of the remaining 1909, 629 completed the survey between June 22 and December 3, 2020, yielding a response rate of 32.9% (American Association for Public Opinion Research (AAPOR) Response Rate #6) and cooperation rate of 56.8% (AAPOR Cooperation Rate #2).^18^

### Measures

At the time of the survey (10-36 weeks post COVID-19 onset) respondents answered the following question: “Have you recovered from COVID-19 to your usual state of health?” Respondents who reported they had recovered provided the length of recovery time. Based on length of recovery time for those who had recovered, and time between COVID-19 onset and survey date for those who had not recovered, we calculated the minimum time to recovery for the entire sample. We defined PASC as persistent symptoms 30+ days (30-day COVID-19) or 60+ days (60-day COVID-19) after COVID-19 onset.

Demographic correlates of interest included age group (18-34, 35-44, 45-54, 55-64, 65+), sex (male, female), race/ethnicity (Hispanic, Non-Hispanic (NH) White, NH Black, NH Other), and annual household income (<$35,000, $35,000-$74,999, $75,000+). Clinical correlates included body mass index (BMI, calculated from self-reported height and weight) and the following pre-existing conditions diagnosed prior to the respondent’s COVID-19 diagnosis: asthma, chronic obstructive pulmonary disease (COPD), hypertension, cardiovascular disease, cerebrovascular disease, diabetes, liver disease, kidney disease, cancer, immunosuppressive condition, autoimmune condition, physical disability, or psychological condition. To examine smoking status, we defined current smokers as respondents who had smoked at least 100 lifetime cigarettes and reported smoking every day or some days immediately prior to their illness. Additionally, we assessed the self-reported severity of symptoms when they were at their worst (mild, moderate, severe, or very severe), and hospitalization or intensive care unit (ICU) stay for COVID-19.

### Analysis

We produced weighted prevalence estimates of PASC overall and by demographic and clinical correlates. We then used modified Poisson regression to examine unadjusted and adjusted prevalence ratios for demographic and clinical correlates of PASC. Additionally, because hospitalization and ICU stay may lead to prolonged recovery regardless of admitting diagnoses,^19,20^ we conducted a sensitivity analysis among non-hospitalized cases in our sample (n=410).

We also produced weighted prevalence estimates of the most common persistent symptoms reported by a subset of 60-day COVID-19 cases (77%) who answered ‘no’ to the question: “Have you recovered from COVID-19 to your usual state of health?” Due to a survey skip pattern, only respondents who had not yet recovered were asked to report the symptoms they were still experiencing at the time of the survey. Since all respondents were at least 60 days post COVID-19 onset at the time of the survey, everyone who specified their persistent symptoms met the criteria for 60-day COVID-19. However, we did not capture persistent symptoms for 30-day or 60-day COVID-19 cases who had recovered by the time they completed the survey.

Missing data were minimal for each covariate (≤ 1.8%), apart from annual household income (11.0%) and PASC (3.9%). We used multiple imputation to impute missing values for all variables apart from the outcomes, resulting in an analytic sample size of 593. All analyses accounted for the complex sampling design using Stata 15.^21^ We set the statistical significance level at p < 0.05 using a 2-sided test.

## Results

Table 1 presents weighted descriptive statistics for the full analytic sample. The majority of respondents were female (56.1%), aged 45 or older (68.2%), and Non-Hispanic (NH) White (46.3%) or NH Black (34.8%). Over half of respondents were obese (53.4%). The most prevalent comorbidities were hypertension (43.0%), diabetes (24.5%), asthma (17.1%) and cardiovascular disease (12.3%). With regard to illness severity, most respondents reported severe (39.5%) or very severe symptoms (26.8%), with about one-third reporting mild (13.3%) or moderate symptoms (20.4%). Nearly one-third were hospitalized (32.4%) and 10.1% were admitted to the ICU during the course of their illness. Among respondents who had not recovered to their usual state of health by the time they completed the survey, all of whom met the criteria for 60-day COVID-19, the most common persistent symptoms were fatigue (52.9%) and shortness of breath (43.9%) (Figure 1).

**Table 1.** **Description of analytic sample (n=593), Michigan COVID-19 Recovery Surveillance Study**

|  | Weighted Percent |
| --- | --- |
| Sex |  |
| Male | 43.9% |
| Female | 56.1% |
| Age group |  |
| 18-34 | 15.1% |
| 35-44 | 16.7% |
| 45-54 | 23.6% |
| 55-64 | 25.1% |
| 65+ | 19.6% |
| Race/ethnicity |  |
| Hispanic | 6.0% |
| Non-Hispanic White | 46.3% |
| Non-Hispanic Black | 34.8% |
| Non-Hispanic Other | 12.9% |
| Annual household income |  |
| <\$35,000 | 31.6% |
| \$35,000 - \$74,999 | 26.8% |
| \$75,000 | 41.6% |
| Current smoker prior to illness | 6.3% |
| Body Mass Index |  |
| Underweight/normal weight (BMI < 25) | 15.0% |
| Overweight (BMI 25 to < 30) | 31.6% |
| Obese (BMI 30+) | 53.4% |
| Comorbidities |  |
| Asthma | 17.1% |
| Chronic obstructive pulmonary disease | 5.6% |
| Hypertension | 43.0% |
| Cardiovascular disease | 12.3% |
| Cerebrovascular disease | 3.8% |
| Diabetes | 24.5% |
| Liver disease | 2.3% |
| Kidney disease | 7.1% |
| Cancer | 8.6% |
| Immunosuppressive condition | 3.0% |
| Autoimmune condition | 6.5% |
| Physical disability | 8.7% |
| Psychological condition | 9.2% |
| Severity of COVID-19 illness |  |
| Self-reported symptom severity |  |
| Mild | 13.3% |
| Moderate | 20.4% |
| Severe | 39.5% |
| Very severe | 26.8% |
| Hospitalized | 32.4% |
| Admitted to Intensive Care Unit | 10.1% |

**Figure 1.**
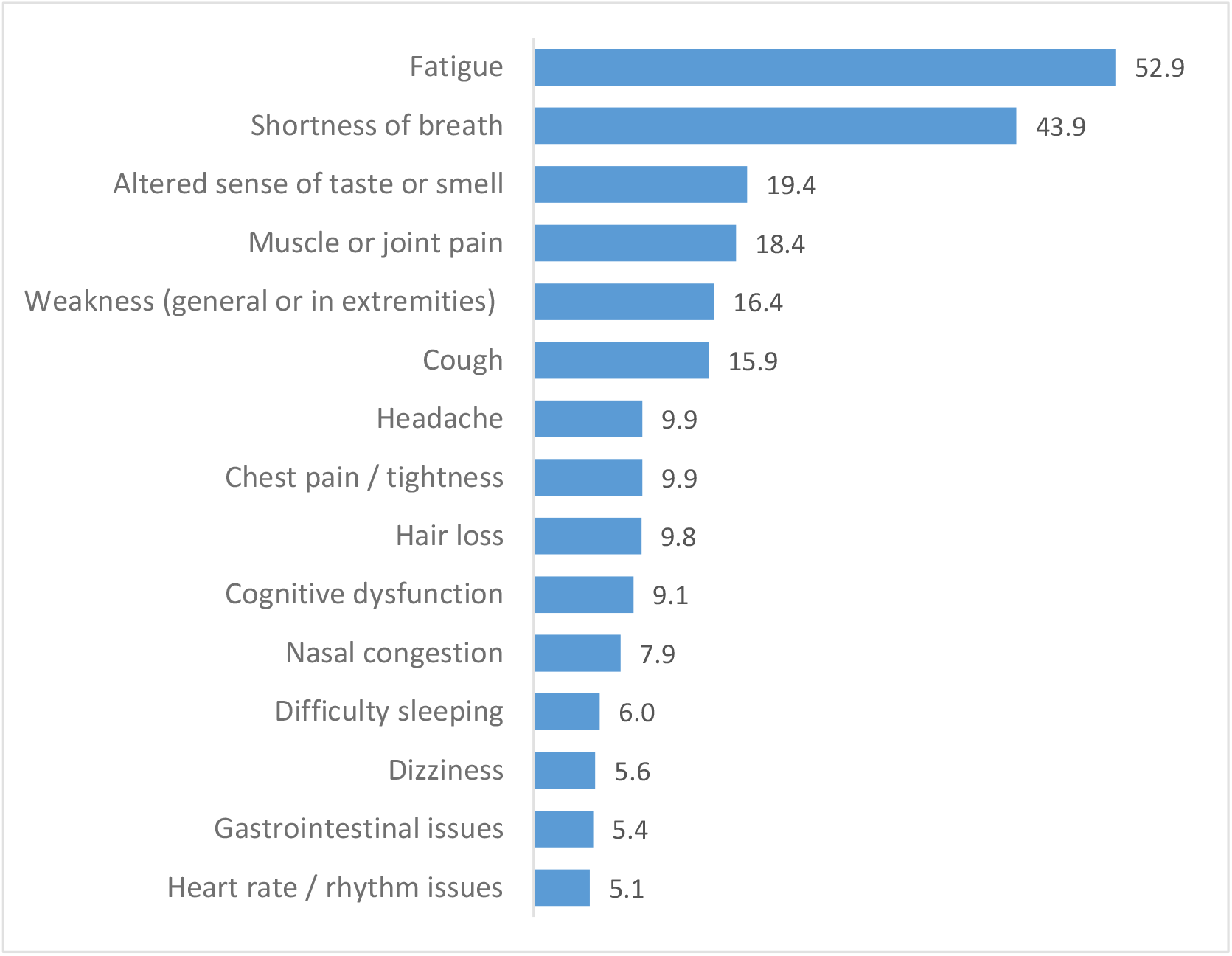
Prevalent symptoms among respondents with 60-Day COVID-19^+^ who had not recovered by the time they were surveyed (n=170)^ +Defined as persistent symptoms at least 60 days post COVID-19 onset ^Respondents who answered ‘no’ to the question: “Have you recovered from COVID-19 to your usual state of health?”

Both 30-day and 60-day COVID-19 were highly prevalent in our sample of symptomatic cases (52.5% and 35.0%, respectively; Table 2). Although the prevalence tended to increase with increasing age and disease severity, 30-day and 60-day COVID-19 were still notably prevalent among respondents who were 18-34 years old (34.9% and 21.2%), reported mild symptoms (29.2% and 24.5%), or did not require hospitalization (43.7% and 26.9%, respectively).

**Table 2.**
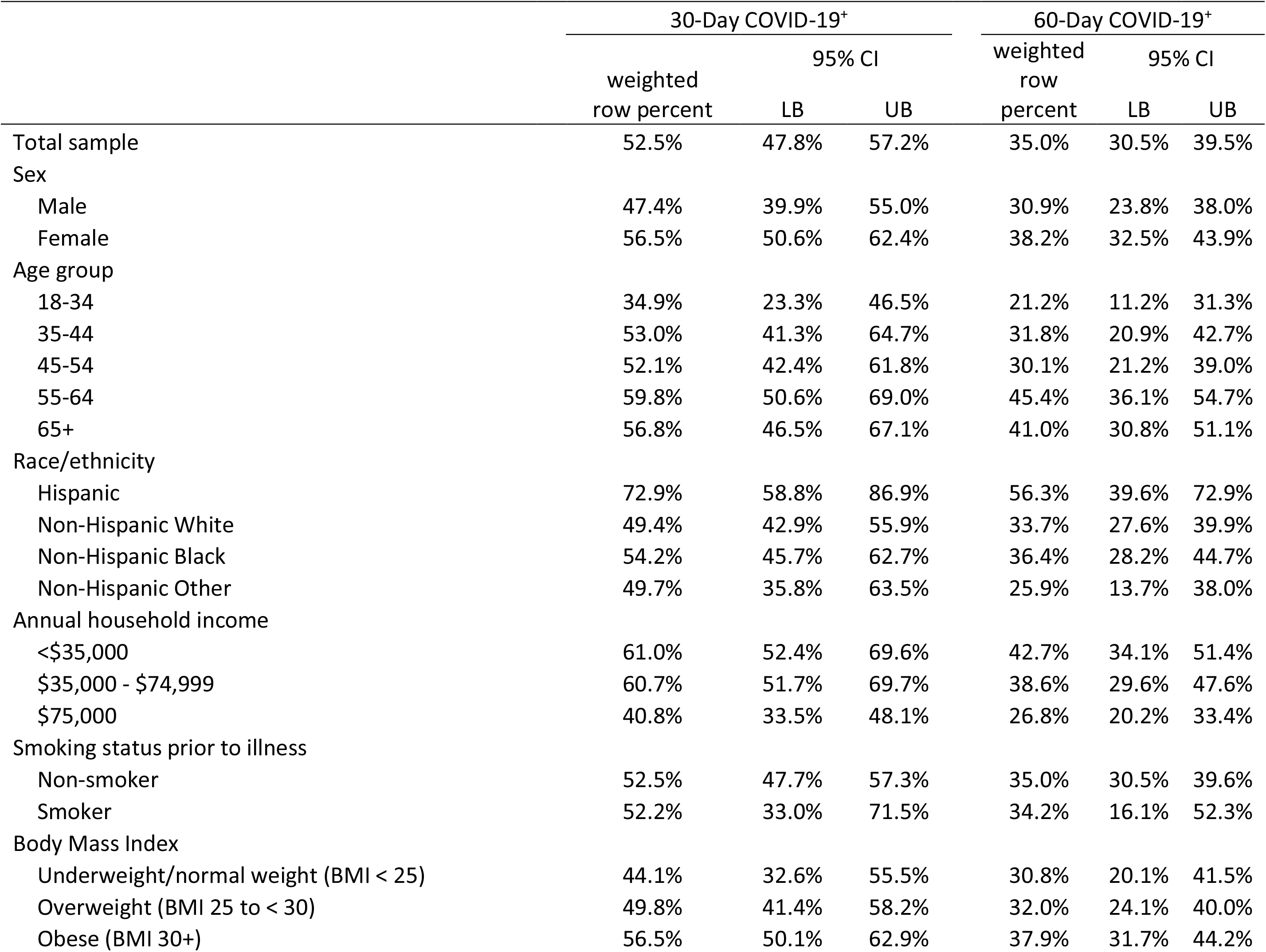
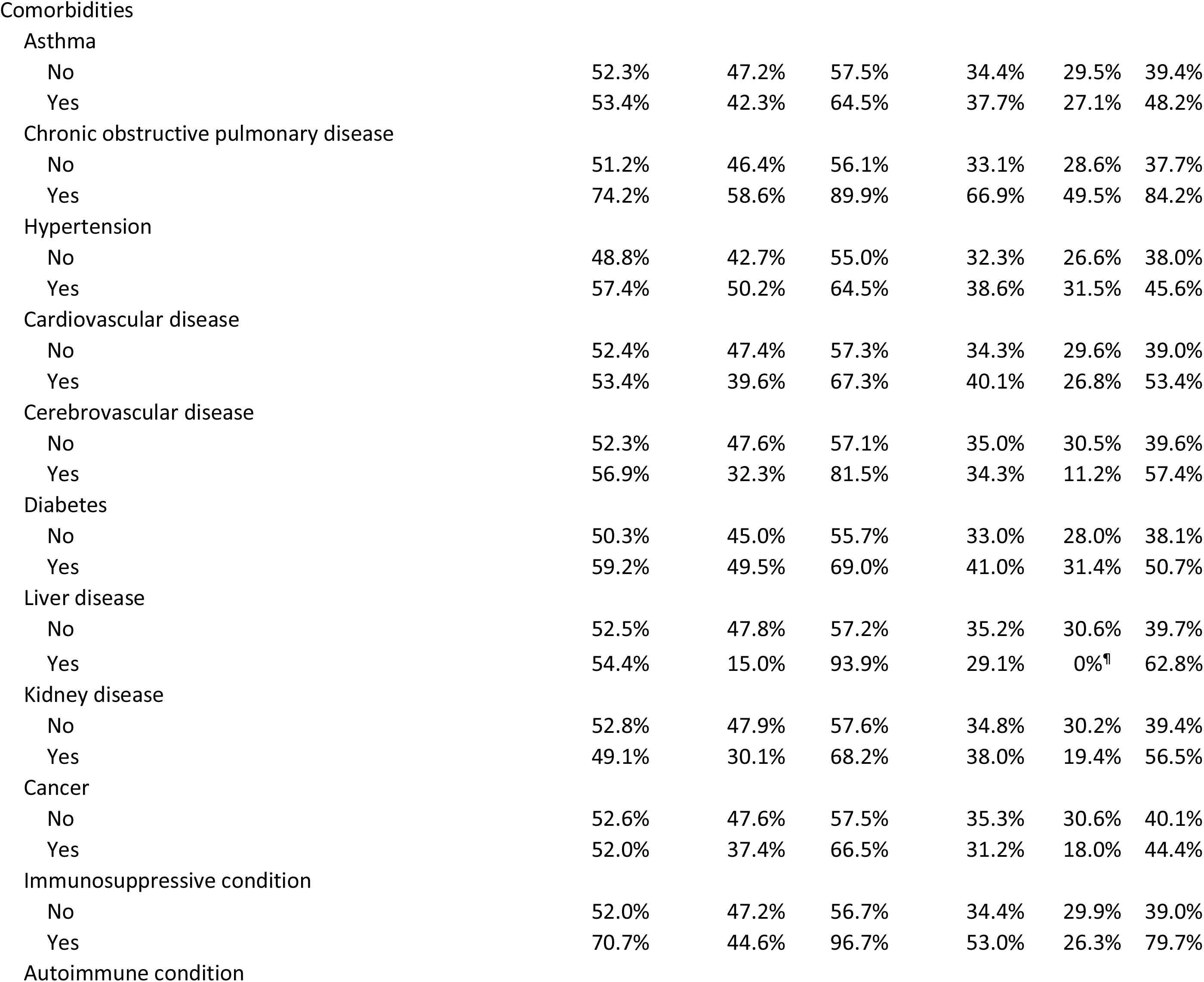
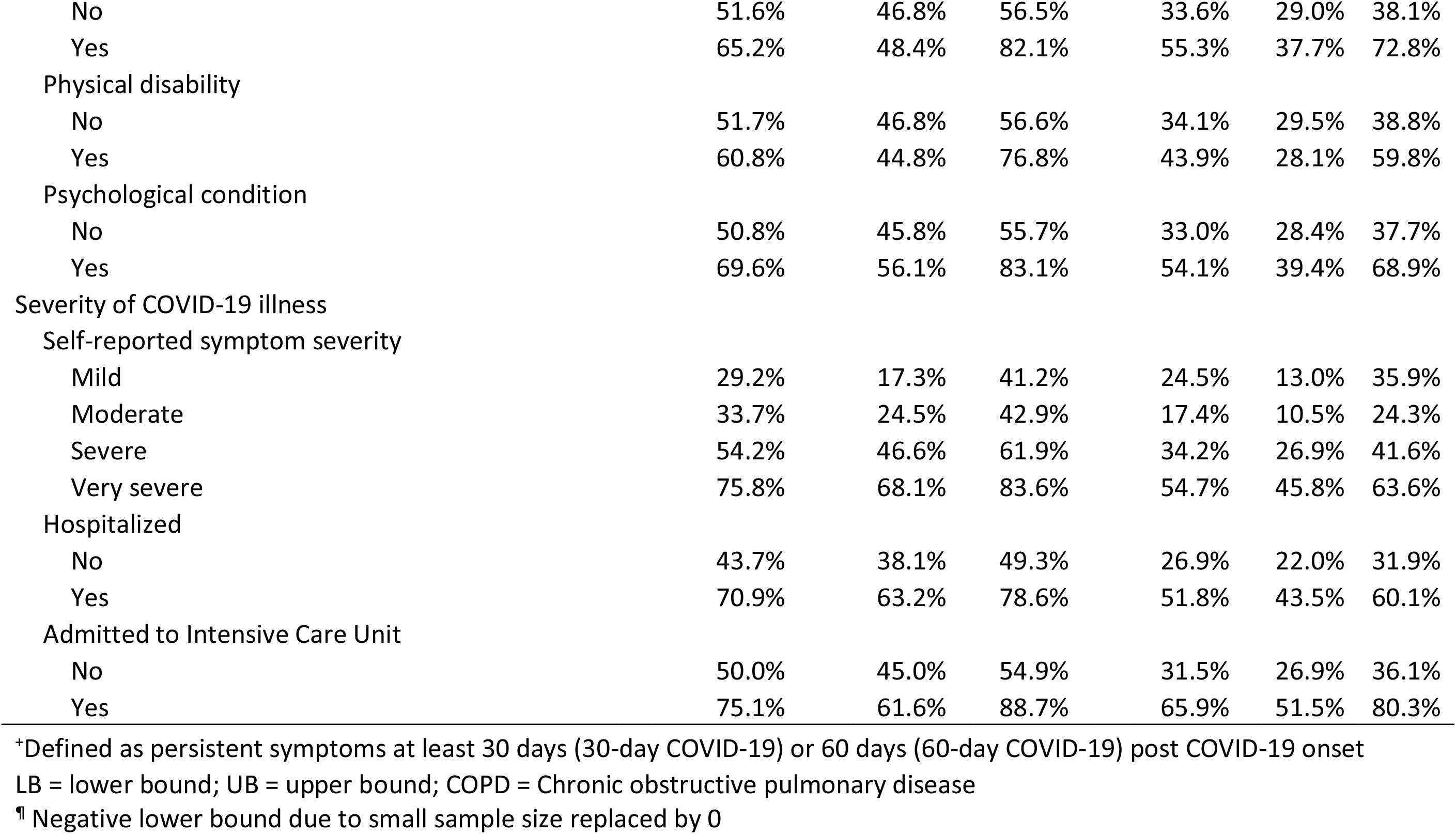
**Prevalence of post-acute sequelae of SARS-CoV-2 infection (PASC) by demographic and clinical factors (n=593), Michigan COVID-19 Recovery Surveillance Study**

|  | 30-Day COVID-19<sup>+</sup> |  |  | 60-Day COVID-19<sup>+</sup> |  |  |
| --- | --- | --- | --- | --- | --- | --- |
|  | weighted<br>row percent | 95% CI |  | weighted<br>row<br>percent | 95% CI |  |
|  |  | LB | UB |  | LB | UB |
| Total sample | 52.5% | 47.8% | 57.2% | 35.0% | 30.5% | 39.5% |
| Sex |  |  |  |  |  |  |
| Male | 47.4% | 39.9% | 55.0% | 30.9% | 23.8% | 38.0% |
| Female | 56.5% | 50.6% | 62.4% | 38.2% | 32.5% | 43.9% |
| Age group |  |  |  |  |  |  |
| 18-34 | 34.9% | 23.3% | 46.5% | 21.2% | 11.2% | 31.3% |
| 35-44 | 53.0% | 41.3% | 64.7% | 31.8% | 20.9% | 42.7% |
| 45-54 | 52.1% | 42.4% | 61.8% | 30.1% | 21.2% | 39.0% |
| 55-64 | 59.8% | 50.6% | 69.0% | 45.4% | 36.1% | 54.7% |
| 65+ | 56.8% | 46.5% | 67.1% | 41.0% | 30.8% | 51.1% |
| Race/ethnicity |  |  |  |  |  |  |
| Hispanic | 72.9% | 58.8% | 86.9% | 56.3% | 39.6% | 72.9% |
| Non-Hispanic White | 49.4% | 42.9% | 55.9% | 33.7% | 27.6% | 39.9% |
| Non-Hispanic Black | 54.2% | 45.7% | 62.7% | 36.4% | 28.2% | 44.7% |
| Non-Hispanic Other | 49.7% | 35.8% | 63.5% | 25.9% | 13.7% | 38.0% |
| Annual household income |  |  |  |  |  |  |
| <\$35,000 | 61.0% | 52.4% | 69.6% | 42.7% | 34.1% | 51.4% |
| \$35,000 - \$74,999 | 60.7% | 51.7% | 69.7% | 38.6% | 29.6% | 47.6% |
| \$75,000 | 40.8% | 33.5% | 48.1% | 26.8% | 20.2% | 33.4% |
| Smoking status prior to illness |  |  |  |  |  |  |
| Non-smoker | 52.5% | 47.7% | 57.3% | 35.0% | 30.5% | 39.6% |
| Smoker | 52.2% | 33.0% | 71.5% | 34.2% | 16.1% | 52.3% |
| Body Mass Index |  |  |  |  |  |  |
| Underweight/normal weight (BMI < 25) | 44.1% | 32.6% | 55.5% | 30.8% | 20.1% | 41.5% |
| Overweight (BMI 25 to < 30) | 49.8% | 41.4% | 58.2% | 32.0% | 24.1% | 40.0% |
| Obese (BMI 30+) | 56.5% | 50.1% | 62.9% | 37.9% | 31.7% | 44.2% |

| Comorbidities |  |  |  |  |  |  |
| --- | --- | --- | --- | --- | --- | --- |
| Asthma |  |  |  |  |  |  |
| No | 52.3% | 47.2% | 57.5% | 34.4% | 29.5% | 39.4% |
| Yes | 53.4% | 42.3% | 64.5% | 37.7% | 27.1% | 48.2% |
| Chronic obstructive pulmonary disease |  |  |  |  |  |  |
| No | 51.2% | 46.4% | 56.1% | 33.1% | 28.6% | 37.7% |
| Yes | 74.2% | 58.6% | 89.9% | 66.9% | 49.5% | 84.2% |
| Hypertension |  |  |  |  |  |  |
| No | 48.8% | 42.7% | 55.0% | 32.3% | 26.6% | 38.0% |
| Yes | 57.4% | 50.2% | 64.5% | 38.6% | 31.5% | 45.6% |
| Cardiovascular disease |  |  |  |  |  |  |
| No | 52.4% | 47.4% | 57.3% | 34.3% | 29.6% | 39.0% |
| Yes | 53.4% | 39.6% | 67.3% | 40.1% | 26.8% | 53.4% |
| Cerebrovascular disease |  |  |  |  |  |  |
| No | 52.3% | 47.6% | 57.1% | 35.0% | 30.5% | 39.6% |
| Yes | 56.9% | 32.3% | 81.5% | 34.3% | 11.2% | 57.4% |
| Diabetes |  |  |  |  |  |  |
| No | 50.3% | 45.0% | 55.7% | 33.0% | 28.0% | 38.1% |
| Yes | 59.2% | 49.5% | 69.0% | 41.0% | 31.4% | 50.7% |
| Liver disease |  |  |  |  |  |  |
| No | 52.5% | 47.8% | 57.2% | 35.2% | 30.6% | 39.7% |
| Yes | 54.4% | 15.0% | 93.9% | 29.1% | 0%<sup>¶</sup> | 62.8% |
| Kidney disease |  |  |  |  |  |  |
| No | 52.8% | 47.9% | 57.6% | 34.8% | 30.2% | 39.4% |
| Yes | 49.1% | 30.1% | 68.2% | 38.0% | 19.4% | 56.5% |
| Cancer |  |  |  |  |  |  |
| No | 52.6% | 47.6% | 57.5% | 35.3% | 30.6% | 40.1% |
| Yes | 52.0% | 37.4% | 66.5% | 31.2% | 18.0% | 44.4% |
| Immunosuppressive condition |  |  |  |  |  |  |
| No | 52.0% | 47.2% | 56.7% | 34.4% | 29.9% | 39.0% |
| Yes | 70.7% | 44.6% | 96.7% | 53.0% | 26.3% | 79.7% |
| Autoimmune condition |  |  |  |  |  |  |
| No | 51.6% | 46.8% | 56.5% | 33.6% | 29.0% | 38.1% |
| Yes | 65.2% | 48.4% | 82.1% | 55.3% | 37.7% | 72.8% |
| Physical disability |  |  |  |  |  |  |
| No | 51.7% | 46.8% | 56.6% | 34.1% | 29.5% | 38.8% |
| Yes | 60.8% | 44.8% | 76.8% | 43.9% | 28.1% | 59.8% |
| Psychological condition |  |  |  |  |  |  |
| No | 50.8% | 45.8% | 55.7% | 33.0% | 28.4% | 37.7% |
| Yes | 69.6% | 56.1% | 83.1% | 54.1% | 39.4% | 68.9% |
| Severity of COVID-19 illness |  |  |  |  |  |  |
| Self-reported symptom severity |  |  |  |  |  |  |
| Mild | 29.2% | 17.3% | 41.2% | 24.5% | 13.0% | 35.9% |
| Moderate | 33.7% | 24.5% | 42.9% | 17.4% | 10.5% | 24.3% |
| Severe | 54.2% | 46.6% | 61.9% | 34.2% | 26.9% | 41.6% |
| Very severe | 75.8% | 68.1% | 83.6% | 54.7% | 45.8% | 63.6% |
| Hospitalized |  |  |  |  |  |  |
| No | 43.7% | 38.1% | 49.3% | 26.9% | 22.0% | 31.9% |
| Yes | 70.9% | 63.2% | 78.6% | 51.8% | 43.5% | 60.1% |
| Admitted to Intensive Care Unit |  |  |  |  |  |  |
| No | 50.0% | 45.0% | 54.9% | 31.5% | 26.9% | 36.1% |
| Yes | 75.1% | 61.6% | 88.7% | 65.9% | 51.5% | 80.3% |

<sup>†</sup>Defined as persistent symptoms at least 30 days (30-day COVID-19) or 60 days (60-day COVID-19) post COVID-19 onset
LB = lower bound; UB = upper bound; COPD = Chronic obstructive pulmonary disease
<sup>¶</sup> Negative lower bound due to small sample size replaced by 0

In unadjusted analyses, older age was statistically significantly associated with 30-day and 60-day COVID-19 prevalence (Table 3). Respondents aged 55-64 years had 1.71 times higher prevalence of 30-day COVID-19 (Prevalence Ratio [PR] 1.71, 95% CI 1.19-2.47) and 2.14 times higher prevalence of 60-day COVID-19 (PR 2.14, 95% CI 1.27-3.59) relative to 18-34 year-olds. Point estimates for respondents aged 65+ were similar to respondents aged 55-64, though slightly lower, potentially due to the competing risk of death. After adjusting for other demographic factors, pre-existing comorbidities, and illness severity, older age (45+ years) was not statistically significantly associated with increased 30-day or 60-day COVID-19. Additionally, although females had a higher prevalence of 30-day and 60-day COVID-19 than males, this difference was not statistically significant.

**Table 3.** **Predictors of post-acute sequelae of SARS-CoV-2 infection (PASC) using modified Poisson regression (n=593), Michigan COVID-19 Recovery Surveillance Study**

|  | 30-Day COVID-19<sup>+</sup> |  |  |  | 60-Day COVID-19<sup>+</sup> |  |  |  |
| --- | --- | --- | --- | --- | --- | --- | --- | --- |
|  | Unadjusted |  | Adjusted<sup>^</sup> |  | Unadjusted |  | Adjusted<sup>^</sup> |  |
|  | Prevalence Ratio | 95% CI | Prevalence Ratio | 95% CI | Prevalence Ratio | 95% CI | Prevalence Ratio | 95% CI |
| Sex |  |  |  |  |  |  |  |  |
| Male | 1.00 |  | 1.00 |  | 1.00 |  | 1.00 |  |
| Female | 1.19 | [0.98, 1.44] | 1.16 | [0.97, 1.40] | 1.24 | [0.94, 1.63] | 1.19 | [0.91, 1.55] |
| Age group |  |  |  |  |  |  |  |  |
| 18-34 | 1.00 |  | 1.00 |  | 1.00 |  | 1.00 |  |
| 35-44 | 1.52* | [1.02, 2.26] | 1.48* | [1.02, 2.14] | 1.50 | [0.83, 2.68] | 1.42 | [0.83, 2.43] |
| 45-54 | 1.49* | [1.02, 2.19] | 1.41 | [0.97, 2.04] | 1.42 | [0.81, 2.48] | 1.33 | [0.78, 2.27] |
| 55-64 | 1.71** | [1.19, 2.47] | 1.37 | [0.95, 1.99] | 2.14** | [1.27, 3.59] | 1.61 | [0.97, 2.68] |
| 65+ | 1.63* | [1.12, 2.38] | 1.34 | [0.93, 1.94] | 1.93* | [1.13, 3.29] | 1.52 | [0.88, 2.62] |
| Race/ethnicity |  |  |  |  |  |  |  |  |
| Hispanic | 1.48** | [1.17, 1.86] | 1.14 | [0.87, 1.49] | 1.67** | [1.18, 2.36] | 1.28 | [0.89, 1.83] |
| Non-Hispanic White | 1.00 |  | 1.00 |  | 1.00 |  | 1.00 |  |
| Non-Hispanic Black | 1.10 | [0.89, 1.35] | 0.92 | [0.74, 1.14] | 1.08 | [0.81, 1.44] | 0.93 | [0.68, 1.27] |
| Non-Hispanic Other | 1.01 | [0.74, 1.37] | 0.97 | [0.73, 1.29] | 0.77 | [0.46, 1.27] | 0.78 | [0.46, 1.31] |
| Annual household income |  |  |  |  |  |  |  |  |
| <\$35,000 | 1.50*** | [1.18, 1.89] | 1.40** | [1.09, 1.79] | 1.59** | [1.15, 2.20] | 1.31 | [0.93, 1.85] |
| \$35,000 - \$74,999 | 1.49*** | [1.18, 1.87] | 1.38** | [1.09, 1.75] | 1.44* | [1.03, 2.02] | 1.27 | [0.90, 1.78] |
| \$75,000 | 1.00 | | 1.00 | | 1.00 | | 1.00 | |
| Current smoker prior to illness | 0.99 | [0.68, 1.45] | 1.18 | [0.82, 1.71] | 0.98 | [0.57, 1.68] | 1.11 | [0.63, 1.95] |
| BMI |  |  |  |  |  |  |  |  |
| underweight/normal weight | 1.00 |  | 1.00 |  | 1.00 |  | 1.00 |  |
| overweight | 1.13 | [0.83, 1.55] | 0.98 | [0.74, 1.29] | 1.04 | [0.68, 1.60] | 0.89 | [0.60, 1.32] |
| obese | 1.28 | [0.96, 1.70] | 0.96 | [0.73, 1.25] | 1.23 | [0.84, 1.81] | 0.93 | [0.63, 1.36] |
| Comorbidities |  |  |  |  |  |  |  |  |
| Asthma | 1.02 | [0.81, 1.28] | 0.92 | [0.74, 1.14] | 1.09 | [0.80, 1.50] | 0.96 | [0.71, 1.30] |
| COPD | 1.45** | [1.15, 1.83] | 1.07 | [0.80, 1.42] | 2.02*** | [1.51, 2.71] | 1.40 | [0.96, 2.02] |
| Hypertension | 1.17 | [0.98, 1.40] | 1.02 | [0.84, 1.24] | 1.19 | [0.93, 1.54] | 1.00 | [0.76, 1.31] |
| Cardiovascular disease | 1.02 | [0.77, 1.34] | 0.96 | [0.72, 1.28] | 1.17 | [0.82, 1.68] | 1.05 | [0.70, 1.57] |
| Cerebrovascular disease | 1.09 | [0.70, 1.69] | 0.97 | [0.65, 1.46] | 0.98 | [0.49, 1.94] | 0.76 | [0.41, 1.41] |
| Diabetes | 1.18 | [0.97, 1.43] | 1.07 | [0.87, 1.32] | 1.24 | [0.94, 1.64] | 1.03 | [0.77, 1.37] |
| Liver disease | 1.02 | [0.49, 2.13] | 0.91 | [0.44, 1.91] | 0.82 | [0.26, 2.61] | 0.66 | [0.26, 1.70] |
| Kidney disease | 0.93 | [0.62, 1.39] | 0.72 | [0.49, 1.06] | 1.09 | [0.66, 1.81] | 0.85 | [0.51, 1.40] |
| Cancer | 0.99 | [0.73, 1.33] | 0.97 | [0.74, 1.26] | 0.88 | [0.57, 1.38] | 0.74 | [0.48, 1.14] |
| Immunosuppressive condition | 1.36 | [0.92, 1.99] | 1.16 | [0.76, 1.78] | 1.53 | [0.91, 2.59] | 1.12 | [0.61, 2.07] |
| Autoimmune condition | 1.26 | [0.96, 1.66] | 1.13 | [0.85, 1.51] | 1.65** | [1.17, 2.32] | 1.46 | [0.98, 2.17] |
| Physical disability | 1.17 | [0.89, 1.56] | 0.87 | [0.64, 1.18] | 1.29 | [0.87, 1.89] | 0.82 | [0.54, 1.24] |
| Psychological condition | 1.37** | [1.10, 1.70] | 1.22 | [0.95, 1.59] | 1.64** | [1.20, 2.23] | 1.42* | [1.00, 2.00] |
| Severity of COVID-19 illness |  |  |  |  |  |  |  |  |
| Self-reported symptom severity |  |  |  |  |  |  |  |  |
| Mild | 1.00 |  |  |  | 1.00 |  | 1.00 |  |
| Moderate | 1.15 | [0.70, 1.89] | 1.25 | [0.78, 2.00] | 0.71 | [0.39, 1.31] | 0.77 | [0.42, 1.42] |
| Severe | 1.86** | [1.20, 2.86] | 1.91** | [1.25, 2.93] | 1.40 | [0.83, 2.35] | 1.36 | [0.81, 2.26] |
| Very severe | 2.59*** | [1.70, 3.95] | 2.25*** | [1.46, 3.46] | 2.24** | [1.36, 3.67] | 1.71* | [1.02, 2.88] |
| Hospitalized | 1.62*** | [1.37, 1.92] | 1.37** | [1.12, 1.69] | 1.92*** | [1.51, 2.45] | 1.40* | [1.02, 1.93] |
| Admitted to Intensive Care Unit | 1.50*** | [1.22, 1.85] | 0.94 | [0.75, 1.18] | 2.09*** | [1.61, 2.72] | 1.16 | [0.84, 1.61] |

+Defined as persistent symptoms at least 30 days (30-day COVID-19) or 60 days (60-day COVID-19) post COVID-19 onset
^Mutually adjusted for all covariates and survey mode
\* p<0.05, \*\* p<0.01, \*\*\* p<0.001
COPD = Chronic obstructive pulmonary disease

Hispanic adults had 48% higher prevalence of 30-day COVID-19 (PR 1.48, 95% CI 1.17-1.86) and 67% higher prevalence of 60-day COVID-19 (PR 1.67, 95% CI 1.18-2.36) than NH White adults in unadjusted models. However, there were no statistically significant differences in 30-day or 60-day COVID-19 by race/ethnicity in the adjusted models. Annual household income was a strong and significant predictor of 30-day COVID-19. Even after adjusting for demographic and clinical factors, respondents with an income less than $75,000 had about 40% higher prevalence of 30-day COVID-19 than respondents with an income at or above $75,000 (<$35,000 aPR 1.40, 95% CI 1.09-1.79; $35,000-74,999 aPR 1.38, 95% CI 1.09-1.75). However, income was not significantly associated with 60-day COVID-19 in fully adjusted models.

While diagnosed COPD, autoimmune condition, and psychological condition were associated with a higher prevalence of 30-day or 60-day COVID-19, only psychological condition remained statistically significant after adjustment. Respondents with (vs. without) a psychological condition had 42% higher prevalence of 60-day COVID-19 (aPR 1.42, 95% CI 1.00-2.00).

Additionally, acute illness severity was strongly associated with both 30-day and 60-day COVID-19. In adjusted models, respondents with self-reported very severe (vs. mild) symptoms had 2.25 times higher prevalence of 30-day COVID-19 (aPR 2.25, 95% CI 1.46-3.46) and 1.71 times higher prevalence of 60-day COVID-19 (aPR 1.71, 95% CI 1.02-2.88). Hospitalized (vs. non-hospitalized) respondents had 37% higher prevalence of 30-day COVID-19 (aPR 1.37, 95% CI 1.12-1.69) and 40% higher prevalence of 60-day COVID-19 (aPR 1.40, 95% CI 1.02-1.93).

Results from the sensitivity analysis restricting the sample to non-hospitalized respondents were largely consistent with results from the primary analysis for 30-day COVID-19, with one exception (Table 4). Although cardiovascular disease was not associated with 30-day COVID-19 among the entire sample, non-hospitalized respondents with (vs. without) cardiovascular disease had 54% higher prevalence of 30-day COVID-19 (aPR 1.54, 95% CI 1.01-2.34). Results from the 60-day COVID-19 sensitivity analysis differed from the primary analysis in several notable ways. Among non-hospitalized respondents, psychological condition was not associated with higher 60-day COVID-19 prevalence, while diagnosed COPD was. Additionally, although non-hospitalized respondents with severe or very severe (vs. mild) symptoms had higher 30-day COVID-19 prevalence, self-reported symptom severity was not associated with 60-day COVID-19 among non-hospitalized respondents.

**Table 4.** **Predictors of post-acute sequelae of SARS-CoV-2 infection (PASC) among respondents who did not require hospitalization for COVID-19 using modified Poisson regression (n=410), Michigan**

|  | 30-Day COVID-19<sup>+</sup> |  | 60-Day COVID-19<sup>+</sup> |  |
| --- | --- | --- | --- | --- |
|  | Adjusted Prevalence Ratio<sup>^</sup> | 95% CI | Adjusted Prevalence Ratio<sup>^</sup> | 95% CI |
| Sex |  |  |  |  |
| Male | 1.00 |  | 1.00 |  |
| Female | 1.29 | [0.97, 1.71] | 1.48 | [0.97, 2.26] |
| Age group |  |  |  |  |
| 18-34 | 1.00 |  | 1.00 |  |
| 35-44 | 1.55* | [1.02, 2.34] | 1.45 | [0.75, 2.78] |
| 45-54 | 1.21 | [0.79, 1.85] | 1.02 | [0.55, 1.92] |
| 55-64 | 1.26 | [0.82, 1.94] | 1.60 | [0.90, 2.86] |
| 65+ | 1.24 | [0.76, 2.04] | 1.72 | [0.84, 3.54] |
| Race/ethnicity |  |  |  |  |
| Hispanic | 1.15 | [0.77, 1.73] | 1.01 | [0.55, 1.84] |
| Non-Hispanic White | 1.00 |  | 1.00 |  |
| Non-Hispanic Black | 0.93 | [0.65, 1.34] | 0.73 | [0.44, 1.21] |
| Non-Hispanic Other | 1.00 | [0.68, 1.48] | 0.82 | [0.43, 1.57] |
| Annual household income |  |  |  |  |
| <\$35,000 | 1.45* | [1.00, 2.08] | 1.43 | [0.85, 2.41] |
| \$35,000 - \$74,999 | 1.56** | [1.13, 2.15] | 1.38 | [0.87, 2.18] |
| \$75,000 | 1.00 | | 1.00 | |
| Current smoker prior to illness | 1.28 | [0.76, 2.16] | 1.45 | [0.67, 3.11] |
| BMI |  |  |  |  |
| underweight/normal weight | 1.00 |  | 1.00 |  |
| overweight | 0.94 | [0.65, 1.36] | 0.85 | [0.50, 1.46] |
| obese | 0.95 | [0.65, 1.38] | 0.92 | [0.53, 1.59] |
| Comorbidities |  |  |  |  |
| Asthma | 0.92 | [0.66, 1.27] | 0.99 | [0.61, 1.62] |
| COPD | 1.38 | [0.91, 2.08] | 1.80* | [1.02, 3.19] |
| Hypertension | 0.95 | [0.70, 1.30] | 0.92 | [0.58, 1.47] |
| Cardiovascular disease | 1.54* | [1.01, 2.34] | 1.82 | [0.95, 3.49] |
| Cerebrovascular disease | 1.04 | [0.48, 2.26] | 0.40 | [0.12, 1.36] |
| Diabetes | 0.97 | [0.68, 1.39] | 1.15 | [0.72, 1.83] |
| Liver disease | 0.46 | [0.10, 2.08] | 0.27 | [0.03, 2.54] |
| Kidney disease | 0.92 | [0.50, 1.69] | 1.05 | [0.44, 2.51] |
| Cancer | 0.68 | [0.41, 1.14] | 0.49 | [0.24, 1.02] |
| Immunosuppressive condition | 1.23 | [0.57, 2.65] | 1.85 | [0.76, 4.47] |
| Autoimmune condition | 1.22 | [0.79, 1.89] | 1.38 | [0.75, 2.54] |
| Physical disability | 0.82 | [0.47, 1.44] | 0.72 | [0.37, 1.43] |
| Psychological condition | 1.32 | [0.94, 1.83] | 1.40 | [0.83, 2.36] |
| Severity of COVID-19 illness |  |  |  |  |
| Self-reported symptom severity |  |  |  |  |
| Mild | 1.00 |  | 1.00 |  |
| Moderate | 1.25 | [0.69, 2.26] | 0.64 | [0.31, 1.29] |
| Severe | 1.85* | [1.04, 3.30] | 1.09 | [0.58, 2.08] |
| Very severe | 2.43** | [1.33, 4.45] | 1.66 | [0.84, 3.29] |

+Defined as persistent symptoms at least 30 days (30-day COVID-19) or 60 days (60-day COVID-19) post COVID-19 onset
^Mutually adjusted for all covariates with prevalence ratio value and survey mode
\* p<0.05, \*\* p<0.01, \*\*\* p<0.001
COPD = Chronic obstructive pulmonary disease

## Discussion

Our study provides prevalence estimates and correlates of PASC using a probability sample of adults who tested positive in Michigan on or before April 15, 2020. Among symptomatic individuals, persistent symptoms were common: 52.5% had not recovered 30 days post COVID-19 onset (30-day COVID-19) and 35.0% had not recovered 60 days post onset (60-day COVID-19). In fully adjusted models, age (35-44 vs. 18-34 only), low income, self-reported severe or very severe (vs. mild) symptoms, and hospitalization statistically significantly predicted 30-day COVID-19, while having a diagnosed psychological disorder, very severe symptoms, and hospitalization statistically significantly predicted 60-day COVID-19.

In the existing literature, we are only aware of one other probability sample among all diagnosed COVID-19 cases in a geographically defined population examining PASC,^13^ with the remaining studies limited to hospitalized,^3,4,7-9^ outpatient,^2,10,12^ or other non-probability samples.^5,6,11^ The one population-based study, conducted in the Faroe Islands among a predominantly non-hospitalized sample, reported 53.1% with at least one symptom an average of 125 days post symptom onset.^13^ While this estimate is nearly identical to our finding of 30-day COVID-19 prevalence (52.5%), it is higher than our estimate of 60-day COVID-19 (35.0%), possibly due to differences in the populations studied.

Although 30-day and 60-day COVID-19 were more prevalent among individuals with severe illness in our study, they were still highly prevalent among individuals with mild to moderate illness. Our finding that 43.7% of non-hospitalized respondents had symptoms 30 days post COVID-19 onset is greater than, but comparable to, a recent study among 14 U.S. academic health systems reporting that 35% of symptomatic outpatient cases had persistent symptoms 14-21 days post SARS-CoV-2 test.^12^ These results add to the growing body of evidence that a sizable proportion of symptomatic COVID-19 cases of varying severity experience PASC. Consistent with recent studies,^11-13^ older age was associated with persistent COVID-19 symptoms. This finding was no longer statistically significant in adjusted models, suggesting the association with age was due to a higher prevalence of comorbidities and severe illness among older age groups. While Hispanic adults had a higher 30- and 60-day COVID-19 prevalence than NH White adults, this relationship was attenuated after adjusting for demographic and clinical factors. We found no statistically significant difference between NH Black and NH White adults in 30- or 60-day COVID-19 prevalence, supporting the growing body of evidence that Black/White inequities in COVID-19 outcomes are primarily driven by increased risk of infection for Black individuals, rather than differences in disease severity among those infected.^22-24^

Apart from psychiatric conditions, comorbidities did not statistically significantly predict 30- or 60-day COVID-19, suggesting another mechanism underlies the risk of prolonged illness.

Moreover, individuals with lower household income (<$75,000) had about 40% higher prevalence of 30-day COVID-19 in fully adjusted models. One potential hypothesis is that greater illness severity, determined by viral dose,^25^ increases the risk of PASC. This pathway could explain the link between low income and post-acute illness, given that low income individuals may be exposed to a greater viral dose due to inability to work from home,^26,27^ lack of adequate personal protective equipment,^28^ and overcrowded living conditions.^26^

There is increasing evidence that illness severity, measured by number of symptoms during the acute phase,^5,11,13^ clinical severity score,^9^ or hospitalization,^11^ may increase the risk of PASC. We found that individuals who reported very severe symptoms (vs. mild) were more than twice as likely to have 30-day COVID-19 and 1.71 times as likely to have 60-day COVID-19. Additionally, individuals requiring hospitalization were about 40% more likely to have 30- or 60-day COVID-19 than non-hospitalized individuals in our study. Severe illness may trigger a hyper-inflammatory immune response,^25,29^ leading to a prolonged recovery period.^30^ Future research is needed to better understand explanatory pathways between illness severity and PASC, including the potential role of viral dose and immune response.

Our study has several limitations. Our sample includes individuals with COVID-19 onset early in the pandemic when access to testing was limited, which may bias our results in two ways. First, if severe cases are overrepresented in our sample, we may be overestimating the prevalence of PASC. Second, given the disproportionate lack of testing access for minority and low-income communities,^31,32^ our sample may underestimate sociodemographic disparities in PASC. Additionally, our data may be subject to both recall and response bias. Because respondents completed interviews between 10-36 weeks post COVID-19 onset, recall bias may be greater for respondents with a longer period between onset and survey completion. Furthermore, individuals with more severe disease or prolonged symptoms may be more likely to participate. Nevertheless, since the sampling frame includes all Michigan residents who tested positive, and symptoms among respondents ranged from mild to very severe, our sample is more comprehensive than previous studies.

## Conclusion

PASC, defined as persistent symptoms 30 or 60 days post onset, is highly prevalent among cases with severe initial symptoms, and, to a lesser extent, cases with mild and moderate symptoms. Formal, coordinated surveillance of post-acute illness is needed to better understand the disease and provide guidance for clinical management. Although we are still attempting to control COVID-19 spread and treat acute illness, we cannot postpone developing robust efforts to characterize and treat PASC, which may potentially affect millions of COVID-19 survivors worldwide.

## Data Availability

The MI CReSS data is not yet available for public download.

## Acknowledgements

This work was supported by funds from the Michigan Department of Health and Human Services, the Michigan Public Health Institute, the University of Michigan Institute for Data Science, and the University of Michigan Rogel Cancer Center.

We would like to thank the Michigan COVID-19 Recovery Surveillance Study participants and interviewers for making this study possible. We would also like to acknowledge Yanmei Xie for her assistance with multiple imputation.

The authors have no conflicts of interest to declare.

